# COVID-19 pandemic dynamics in Ukraine after September 1, 2020

**DOI:** 10.1101/2020.12.21.20248627

**Authors:** Igor Nesteruk

**Affiliations:** Institute of Hydromechanics, National Academy of Sciences of Ukraine National Technical University of Ukraine “Igor Sikorsky Kyiv Polytechnic Institute”.

**Keywords:** COVID-19 pandemic, epidemic dynamics in Ukraine, mathematical modeling of infection diseases, SIR model, parameter identification, statistical methods

## Abstract

**Background:** The threats of the COVID-19 pandemic require the mobilization of scientists, including mathematicians. To understand how the number of cases increases versus time, various models based on direct observations of a random number of new cases and differential equations can be used. Complex mathematical models contain many unknown parameters, the values of which must be determined using a limited number of observations of the disease over time. Even long-term monitoring of the epidemic may not provide reliable estimates of its parameters due to the constant change of testing conditions, isolation of infected and quarantine. Therefore, simpler approaches should also be used, for example, some smoothing of the dependence of the number of cases on time and the known SIR (susceptible-infected-removed) model. These approaches allowed to detect the waves of pandemic in different countries and regions and to make adequate predictions of the duration, hidden periods, reproduction numbers, and final sizes of its waves. In particular, seven waves of the COVID-19 pandemic in Ukraine were investigated.

**Objective:** We will detect new epidemic waves in Ukraine that occurred after September 1, 2020 and estimate the epidemic characteristics with the use of generalized SIR model. Some predictions of the epidemic dynamics will be presented.

**Methods:** In this study we use the smoothing method for the dependence of the number of cases on time; the generalized SIR model for the dynamics of any epidemic wave, the exact solution of the linear differential equations and statistical approach developed before.

**Results:** Seventh and eights epidemic waves in Ukraine were detected and the reasons of their appearance were discussed. The optimal values of the SIR model parameters were calculated. The prediction for the COVID-19 epidemic dynamics in Ukraine is not very optimistic: new cases will not stop appearing until June 2021. Only mass vaccination and social distancing can change this trend.

**Conclusions:** New waves of COVID-19 pandemic can be detected, calculated and predicted with the use of rather simple mathematical simulations. The expected long duration of the pandemic forces us to be careful and in solidarity.The government and all Ukrainians must strictly adhere to quarantine measures in order to avoid fatal consequences.

## Introduction

The studies of the COVID-19 pandemic dynamics in Ukraine are presented in [1-14] and summarized in the book [15]. Different simulation and comparison methods together with the official WHO data sets, [16] were used. In particular the classical SIR model [17-19], connecting the number of susceptible *S*, infected and spreading the infection *I* and removed *R* persons, was applied in [2-5, 8-10] to simulate the first pandemic wave in Ukraine. The unknown parameters of this model were be estimated with the use of the cumulative number of cases *V=I+R* and the statistics-based method of parameter identification developed in [20, 21].

The weakening of quarantine restrictions, changes in the social behavior and probably in the coronavirus activity causes change in SIR characteristics and the epidemic dynamics. To detect these changes, a simple method of numerical differentiations of accumulated number of cases was proposed in [11, 15]. To simulate these new pandemic waves, the SIR model was generalized in [12, 15]. In [12, 14, 15] the results of simulation of the first six epidemic waves in Ukraine are presented with the use of a procedure for sequentially determining the parameters of the model for each epidemic wave, starting with the first one.

This method requires considerable effort and time. The book [15] introduced a new algorithm for determining the optimal parameter values for a particular epidemic wave without calculating the dynamics of previous waves and presented calculations for the seventh epidemic wave in Ukraine. In this paper, we will analyze the dynamics of the epidemic in Ukraine in the period from September 1 to December 20, 2020, calculate the parameters of the eighth wave and make some predictions.

## Materials and Methods

### Data

The official information regarding the accumulated numbers of confirmed COVID-19 cases *V*_*j*_ in Ukraine according to the official sources [22, 23] is shown in Table 1. Unfortunately, WHO stopped to present the daily number of new cases in August 2020. The corresponding moments of time *t*_*j*_ (measured in days, zero point is January 20, 2020) are also shown in this table. To calculate the SIR characteristics for the seventh epidemic wave in Ukraine, the data set for the period October 1-14 was used in [15]. We will use the period November 21 – December 4, 2020 to calculate the characteristics of the eighth epidemic wave. Other values presented in Table 1 were used only for comparisons and verifications of calculations.

**Table 1.**
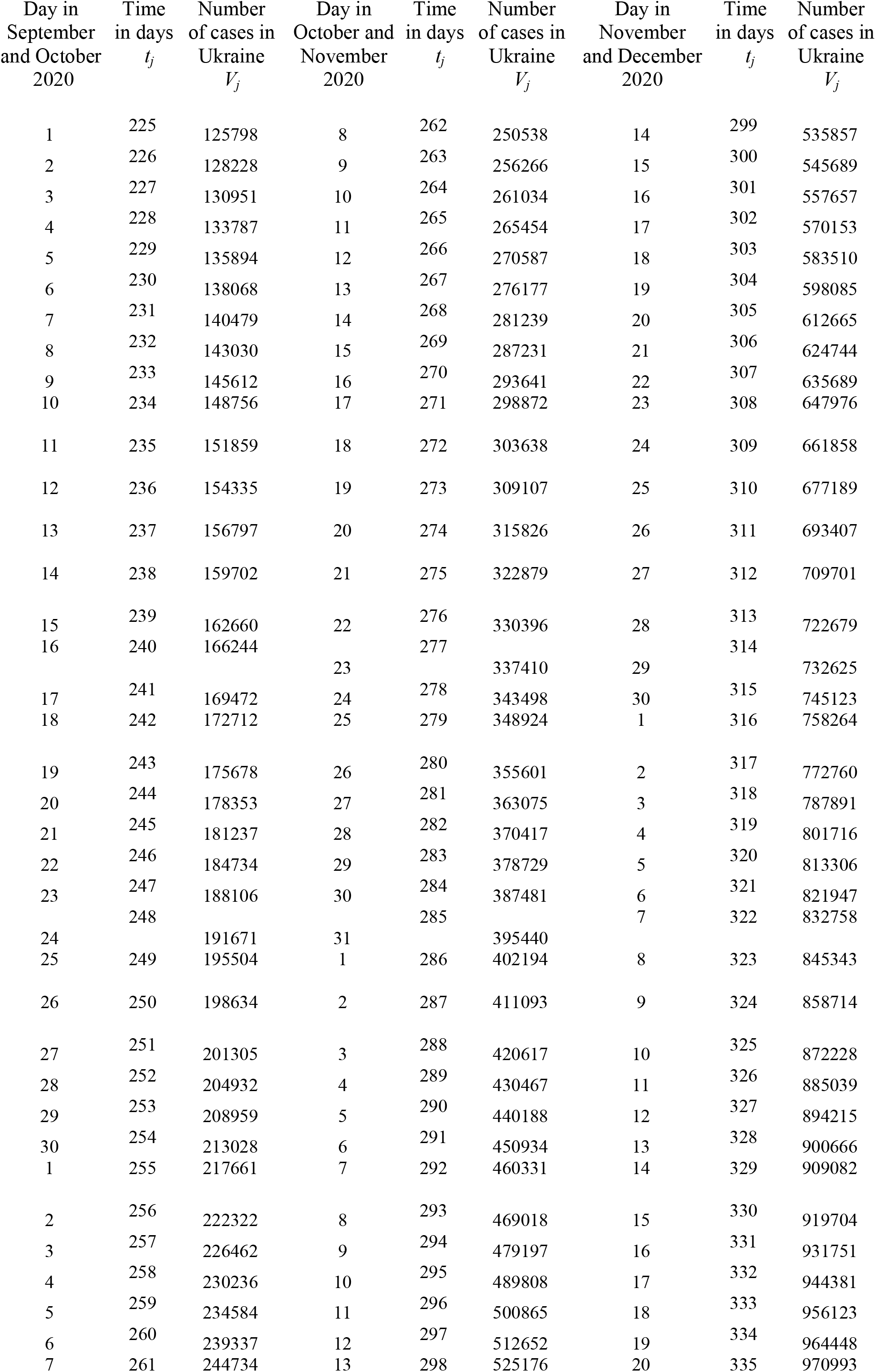
Official cumulative numbers of confirmed Covid-19 cases in Ukraine, [22, 23].

| Day in<br>September<br>and October<br>2020 | Time<br>in days<br>$t_j$ | Number<br>of cases in<br>Ukraine<br>$V_j$ | Day in<br>October and<br>November<br>2020 | Time<br>in days<br>$t_j$ | Number<br>of cases in<br>Ukraine<br>$V_j$ | Day in<br>November<br>and December<br>2020 | Time<br>in days<br>$t_j$ | Number<br>of cases in<br>Ukraine<br>$V_j$ |
| --- | --- | --- | --- | --- | --- | --- | --- | --- |
| 1 | 225 | 125798 | 8 | 262 | 250538 | 14 | 299 | 535857 |
| 2 | 226 | 128228 | 9 | 263 | 256266 | 15 | 300 | 545689 |
| 3 | 227 | 130951 | 10 | 264 | 261034 | 16 | 301 | 557657 |
| 4 | 228 | 133787 | 11 | 265 | 265454 | 17 | 302 | 570153 |
| 5 | 229 | 135894 | 12 | 266 | 270587 | 18 | 303 | 583510 |
| 6 | 230 | 138068 | 13 | 267 | 276177 | 19 | 304 | 598085 |
| 7 | 231 | 140479 | 14 | 268 | 281239 | 20 | 305 | 612665 |
| 8 | 232 | 143030 | 15 | 269 | 287231 | 21 | 306 | 624744 |
| 9 | 233 | 145612 | 16 | 270 | 293641 | 22 | 307 | 635689 |
| 10 | 234 | 148756 | 17 | 271 | 298872 | 23 | 308 | 647976 |
| 11 | 235 | 151859 | 18 | 272 | 303638 | 24 | 309 | 661858 |
| 12 | 236 | 154335 | 19 | 273 | 309107 | 25 | 310 | 677189 |
| 13 | 237 | 156797 | 20 | 274 | 315826 | 26 | 311 | 693407 |
| 14 | 238 | 159702 | 21 | 275 | 322879 | 27 | 312 | 709701 |
| 15 | 239 | 162660 | 22 | 276 | 330396 | 28 | 313 | 722679 |
| 16 | 240 | 166244 | 23 | 277 | 337410 | 29 | 314 | 732625 |
| 17 | 241 | 169472 | 24 | 278 | 343498 | 30 | 315 | 745123 |
| 18 | 242 | 172712 | 25 | 279 | 348924 | 1 | 316 | 758264 |
| 19 | 243 | 175678 | 26 | 280 | 355601 | 2 | 317 | 772760 |
| 20 | 244 | 178353 | 27 | 281 | 363075 | 3 | 318 | 787891 |
| 21 | 245 | 181237 | 28 | 282 | 370417 | 4 | 319 | 801716 |
| 22 | 246 | 184734 | 29 | 283 | 378729 | 5 | 320 | 813306 |
| 23 | 247 | 188106 | 30 | 284 | 387481 | 6 | 321 | 821947 |
| 24 | 248 | 191671 | 31 | 285 | 395440 | 7 | 322 | 832758 |
| 25 | 249 | 195504 | 1 | 286 | 402194 | 8 | 323 | 845343 |
| 26 | 250 | 198634 | 2 | 287 | 411093 | 9 | 324 | 858714 |
| 27 | 251 | 201305 | 3 | 288 | 420617 | 10 | 325 | 872228 |
| 28 | 252 | 204932 | 4 | 289 | 430467 | 11 | 326 | 885039 |
| 29 | 253 | 208959 | 5 | 290 | 440188 | 12 | 327 | 894215 |
| 30 | 254 | 213028 | 6 | 291 | 450934 | 13 | 328 | 900666 |
| 1 | 255 | 217661 | 7 | 292 | 460331 | 14 | 329 | 909082 |
| 2 | 256 | 222322 | 8 | 293 | 469018 | 15 | 330 | 919704 |
| 3 | 257 | 226462 | 9 | 294 | 479197 | 16 | 331 | 931751 |
| 4 | 258 | 230236 | 10 | 295 | 489808 | 17 | 332 | 944381 |
| 5 | 259 | 234584 | 11 | 296 | 500865 | 18 | 333 | 956123 |
| 6 | 260 | 239337 | 12 | 297 | 512652 | 19 | 334 | 964448 |
| 7 | 261 | 244734 | 13 | 298 | 525176 | 20 | 335 | 970993 |

It must be noted that the data presented in Table 1 does not show all the COVID-19 cases in Ukraine. Many infected persons are not identified, since they have no symptoms. Many people know that they are ill, since they have similar symptoms as other members of families, but avoid to make tests. Unfortunately, one laboratory confirmed case can correspond to several other cases which are not confirmed and displayed in the official statistics. This fact reduces the accuracy of mathematical simulations.

### Detection of epidemic waves

To control the changes of epidemic parameters, we can use daily numbers of new cases and their derivatives. Since these values are random, we need some smoothing. For example, we can use the smoothed daily number of accumulated cases proposed in [11, 12, 14, 15]:

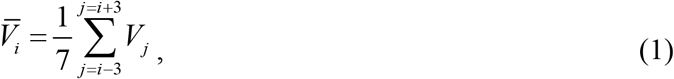

The first and second derivatives can be estimated with the use of following formulas:

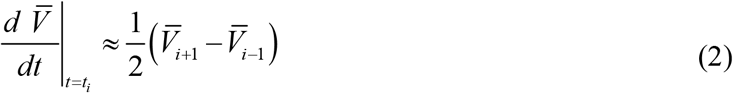

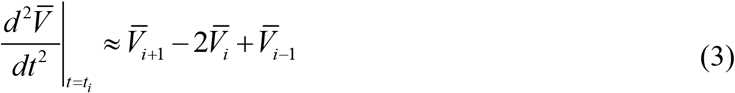

### Generalized SIR model

The classical SIR model for an infectious disease [17-19] was generalized in [12, 15] in order to simulate different epidemic waves. We suppose that the SIR model parameters are constant for every epidemic wave, i.e. for the time periods: 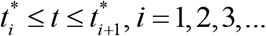. Than for every wave we can use the equations, similar to [17-19]:

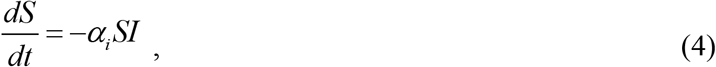

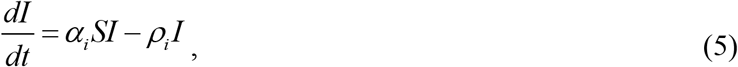

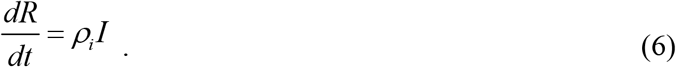

Here *S* is the number of susceptible persons (who are sensitive to the pathogen and **not protected**); *I* is the number of infected persons (who are sick and **spread the infection**; please don’t confuse with the number of still ill persons, so known active cases) and *R* is the number of removed persons (who **no longer spread the infection**; this number is the sum of isolated, recovered, dead, and infected people who left the region). Parameters *α*_*i*_ and *ρ*_*i*_ are supposed to be constant for every epidemic wave.

Parameters *α*_*i*_ show how quick the susceptible persons become infected (see (4)). Large values of this parameter correspond to severe epidemics with many victims. These parameters accumulate many characteristics. First they shows how strong (virulent) is the pathogen and what is the way of its spreading. Parameters *α*_*i*_ accumulate also the frequency of contacts and the way of contacting. In order to decrease the values of *α*_*i*_, we have to minimize the number of our contacts and change our contacting habits. For example, we have to avoid the public places and use masks there, minimize or cancel traveling. We have to change our contact habits: to avoid handshakes and kisses. First, all these simple things are very useful to protect yourself. In addition, if most people follow these recommendations, we have chance to diminish the values of parameters *α*_*i*_ and reduce the negative effects of the pandemic.

The parameters *ρ*_*i*_ characterize the patient removal rates, since eq. (6) demonstrates the increase rate of *R*. The inverse values 1/ *ρ*_*i*_ are the estimations for time of spreading infection *τ*_*i*_ during *i-th* epidemic wave. So, we are interested in increasing the values of parameters *ρ*_*i*_ and decreasing 1/ *ρ*_*i*_. People and public authorities should work on this and organize immediate isolation of suspicious cases.

Since the derivative *d* (*S* + *I* + *R*) / *dt* is equal to zero (it follows from summarizing Eqs. (4)-(6)), the sum

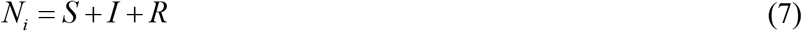

must be constant for every wave and is not the volume of population.

To determine the initial conditions for the set of equations (4)–(6), let us suppose that at the beginning of every epidemic wave 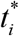:

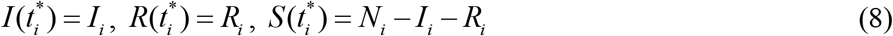

It follows from (4) and (5) that

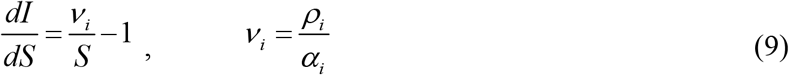

Integration of (9) with the initial conditions (8) yields:

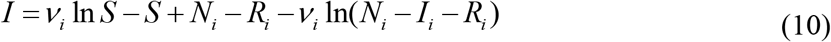

It follows from (9) that function *I* has a maximum at *S* =*v*_*i*_ and tends to zero at infinity. The corresponding number of susceptible persons at infinity *S*_*i*∞_ > 0 can be calculated from a non-linear equation

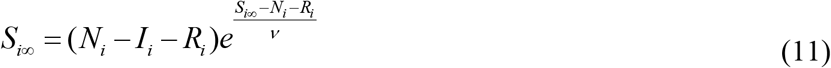

Formula (11) follows from (10) at *I*=0.

In [12, 15] the set of differential equations (4)-(7) was solved by introducing the function

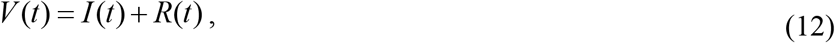

corresponding to the number of victims or the cumulative confirmed number of cases. For many epidemics (including the COVID-9 pandemic) we cannot observe dependencies *S*(*t*), *I* (*t*) and *R*(*t*) but observations of the accumulated number of cases *V*_*j*_ corresponding to the moments of time *t*_*j*_ provide information for direct assessments of the dependence *V* (*t*).

It follows from (5) and (6) that:

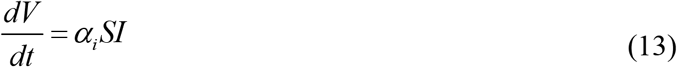

Eqs. (7), (10) and (13) yield:

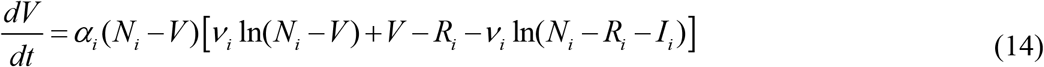

Integration of (14) provides an analytical solution for the set of equations (4)–(6):

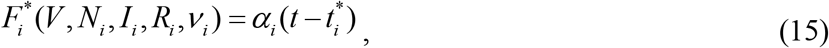

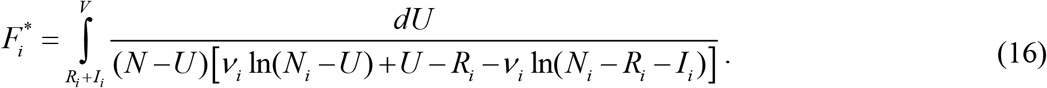

Thus, for every set of parameters *N*_*i*_, *I*_*i*_, *R*_*i*_, *v*_*i*_, *α*_*i*_ and a fixed value of *V*, integral (16) can be calculated and the corresponding moment of time can be determined from (15). Then functions *I(t)* and *R(t)* can be easily calculated with the use of formulas (10) and:

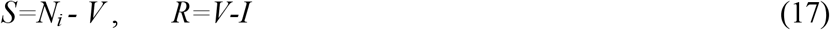

The final numbers of victims (final accumulated number of cases corresponding to the *i-th* epidemic wave) can be calculated from:

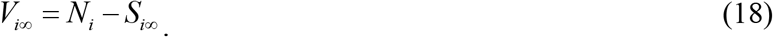

To estimate the final day of the *i-th* epidemic wave, we can use the condition:

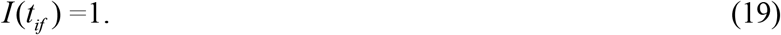

which means that at *t* > *t*_*if*_ less than one person still spreads the infection.

### Parameter identification procedure

In the case of a new epidemic, the values of its parameters are unknown and must be identified with the use of limited data sets. For the first wave of an epidemic starting with one infected person, the number of unknown parameters is only four, since *I*_1_ = 1 and *R*_1_ = 0. The corresponding statistical approach was proposed in [20, 21] and used [2-5, 8-10] to simulate the first COVID-19 pandemic wave in Ukraine.

For the next epidemic waves (*i* > 1), the moments of time 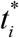 corresponding to their beginning are known. Therefore the exact solution (15)-(16) depend only on five parameters - *N*_*i*_, *I*_*i*_, *R*_*i*_, *v*_*i*_, *α*_*i*_. Then the registered number of victims *V*_*j*_ corresponding to the moments of time *t*_*j*_ can be used in eq. (16) in order to calculate 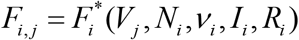 for every fixed values of *N*_*i*_,*v*_*i*_, *I*_*i*_, *R*_*i*_ and then to check how the registered points fit the straight line (15).

Eq. (15) can be rewritten as follows:

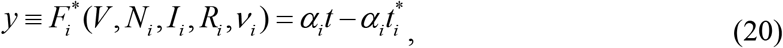

Assuming

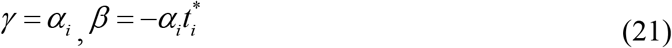

we can estimate the values of parameters *γ* and *β*, by treating the values 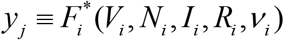 and corresponding time moments *t*_*j*_ as random variables Then we can use the observations of the accumulated number of cases and the linear regression in order to calculate the coefficients 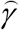 and 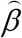 of the regression line

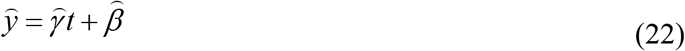

using the standard formulas from, e.g., [24]. Values 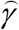 and 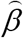 can be treated as statistics-based estimations of parameters *γ* and *β* from relationships (21).

The reliability of the method can be checked by calculating the correlation coefficients *r*_*i*_ (see e.g., [24]) for every epidemic wave checking how close its value is to unity. We can use also the F-test for the null hypothesis that says that the proposed linear relationship (20) fits the data set. The experimental values of the Fisher function can be calculated for every epidemic wave with the use of the formula:

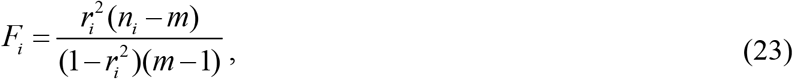

where *n*_*i*_ is the number of observations for the *i*-th epidemic wave, *m* = 2 is the number of parameters in the regression equation. The corresponding experimental value *F*_*i*_ has to be compared with the critical value *F*_*C*_ (*k*_1_, *k*_2_) of the Fisher function at a desired significance or confidence level (*k*_1_ = *m −*1, *k*_2_ = *n*_*i*_ *− m*). When the values *n*_*i*_ and *m* are fixed, the maximum of the Fisher function coincides with the maximum of the correlation coefficient. Therefore, to find the optimal values of parameters *N*_*i*_, *v*_*i*_, *I*_*i*_, *R*_*i*_, we have to find the maximum of the correlation coefficient for the linear dependence (20). To compare the reliability of different predictions (with different values of *n*_*i*_) it is useful to use the ratio *F*_*i*_ / *F*_*C*_ (1, *n*_*i*_ *−* 2) at fixed significance level. We will use the level 0.001; corresponding values of *F*_*C*_ (1, *n*_*i*_ *−* 2) can be taken from [25]. The most reliable prediction yields the highest *F*_*i*_ / *F*_*C*_ (1, *n*_*i*_ *−* 2) ratio.

The exact solution (15)-(16) allows avoiding numerical solutions of differential equations (4)-(6) and significantly reduce the time spent on calculations. In the case of sequential calculation of epidemic waves *i* = 1,2,3 …, it is possible to avoid determining the four optimal unknown parameters *N*_*i*_, *v*_*i*_, *I*_*i*_, *R*_*i*_, thereby reducing the amount of calculations and difficulties in isolation a maximum of the correlation coefficient. For parameters *I*_*i*_, *R*_*i*_ it is possible to use the numbers of *I* and *R* calculated for the previous wave of epidemic at the moment of time when the following wave began. Then we need to calculate values 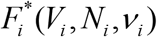, linear regression coefficients (22), correlation coefficient *r*_*i*_, *F*_*i*_ / *F*_*C*_ (1, *n −* 2) and to isolate the values of parameters *N*_*i*_ and *v* _*I*_ corresponding to the maximum of *r*_*i*_. Knowing the optimal values of five parameters *N*_*i*_, *I*_*i*_, *R*_*i*_, *v*_*i*_, *α*_*i*_, the SIR curves and other characteristics of the corresponding epidemic wave can be calculated with the use of formulas (10)-(17). This approach has been successfully used in [12, 14, 15]. In particular, six waves of the Covid-19 epidemic in Ukraine and four pandemic waves in the world were calculated.

Segmentation of epidemic waves and their sequential SIR simulations need a lot of efforts. To avoid this, a new method of obtaining the optimal values of SIR parameters was proposed in [15]. First of all we can use the relationship

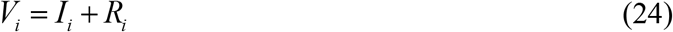

which follows from (12). To estimate the value *V*_*i*_, we can use the smoothed accumulated number of cases (e.g., formula (1)). Then

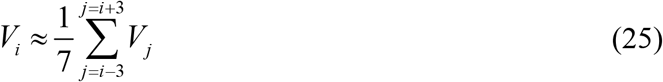

where *i* corresponds to the moment of time 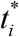. To obtain one more relationship, let us use (7) and (13)

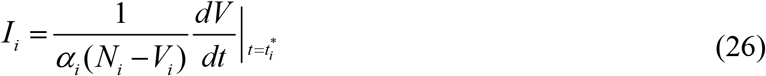

To estimate the average number of new cases *dV/dt* at the moment of time 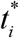, we can use (2). Thus we have only two independent parameters *N*_*i*_ and *v*_*i*_. To calculate the value of parameter *α*_*i*_, some iterations can be used (see details in [15]).

## Results and discussion

The COVID-19 pandemic characteristics for Ukraine in autumn 2020 are shown in Fig. 1. Differentiation of the smoothed number of accumulated cases (eq. (1), line) with the use of formulas (2) (“triangles”) and (3) (stars) allow us to detect the changes in epidemic dynamics. It can be seen that after November 15 the average daily number of new cases (“triangles”) started to decrease. Similar short periods of the epidemic stabilization occurred in May, June and August, 2020 (see [11, 12, 14, 15]). Let us hope that the Christmas and New Year celebrations will not significantly worsen the existing positive dynamics.

**Fig. 1.**
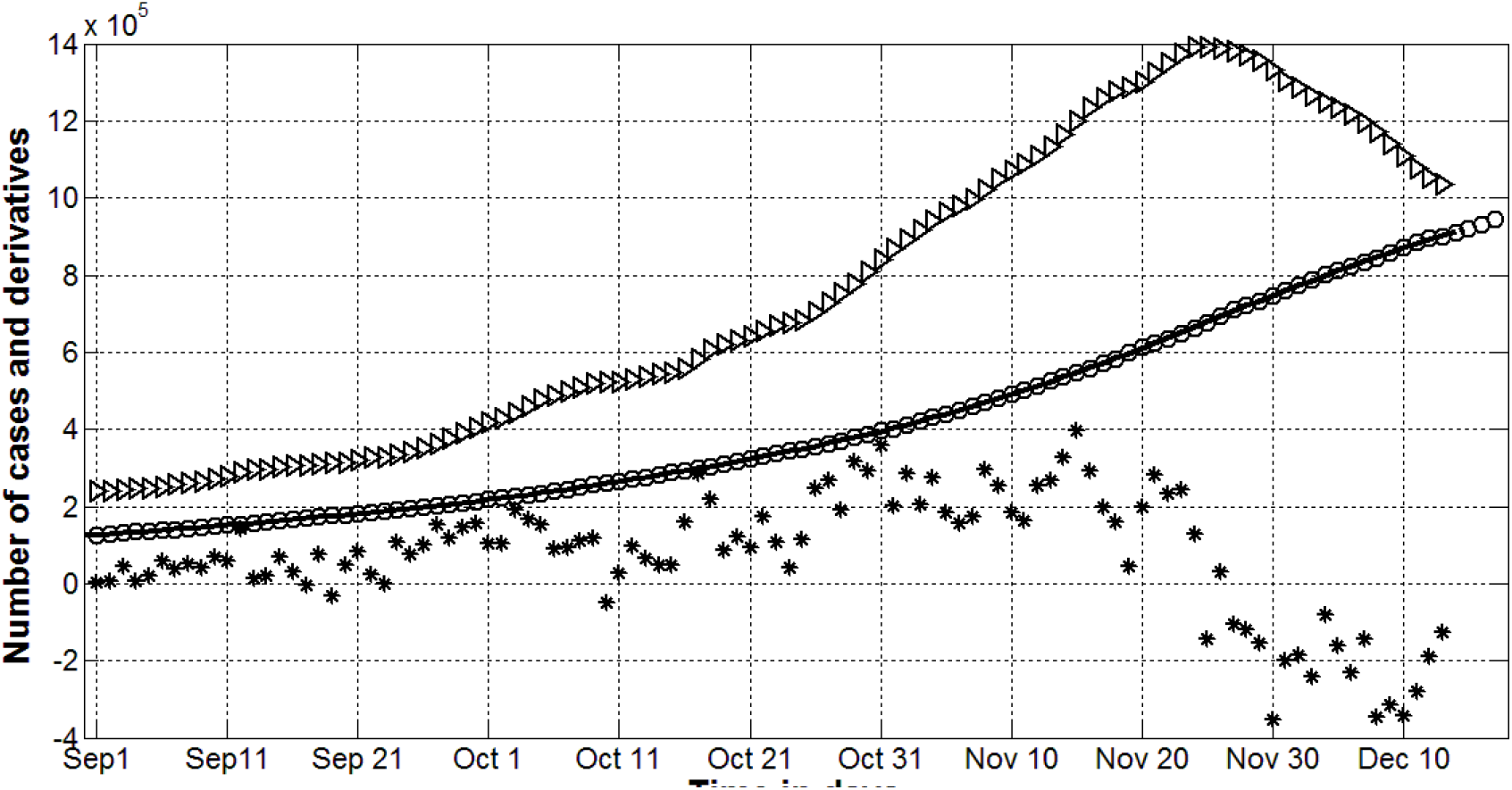
Pandemic dynamics in Ukraine after September 1, 2020. Accumulated number of cases (V_j_ -”cicrles”, Table 1; smoothed values - line, eq. (1)). “Triangles” show the first derivative (eq. (2)) multiplied by 100, “stars” - the second derivative (eq. (3)) multiplied by 1000.

The values of the second derivative (3) allows to detect the changes in the epidemic characteristics and to separate its different waves. The jump in *d*^*2*^*V/dt*^*2*^ values corresponding to September 11 (see “stars” in Fig. 1) can be explained by the beginning of classes in schools and universities (on September 1). Children and young people are often asymptomatic carriers of the infection and bring it to their families. For example, employees of two kindergartens and two schools in the Ukrainian city of Chmelnytskii were tested for antibodies to COVID-19, [26]. In total 292 people work in the surveyed institutions. Some of the staff had already fallen ill with COVID-19 or were hospitalized. Therefore, they were not tested accordingly. Of the 241 educators tested, antibodies were detected in 148, or 61.4%. These results indicate the important role of children in the spread of COVID-19 infection and the fact that in Ukraine those people who have become ill and have antibodies to coronavirus infection, obviously, are much more than in the official recovery statistics presented in Table 1.

The severe jumps in *d*^*2*^*V/dt*^*2*^ values occurred also in October and November, 2020 (see “stars” in Fig. 1). Probably, this is due to the local elections and a presidential poll, which were held throughout Ukraine on October 25, 2020 and involved hundreds of thousands of people to campaign and work in election commissions (their number was about 30 thousand). This obviously increased the number of contacts and the likelihood of additional infections. The corresponding seventh epidemic wave in Ukraine was considered in [15] (the results for the first six waves can be found in [12, 14, 15]). The optimal values of SIR parameters; predictions of the epidemic wave duration *t*_*if*_ and its final size *V*_*i*∞_ are shown in Table 2. The characteristics of the eights wave (occurred after November 21, 2020) were calculated and presented in Table 2. It can be seen that the predictions for Ukraine are very pessimistic. The number of cases will exceed 1.07 million and new cases will appear even in June 2021.

**Table 2.** Calculated optimal values of SIR parameters for the sixth, seventh and eighth waves of the COVID-19 epidemic in Ukraine

| Characteristics | Sixth wave,<br>I=6, n=14, [14, 15] | Seventh wave,<br>i=7, n=14, [15] | Eighth wave,<br>i=8, n=14 |
| --- | --- | --- | --- |
| Period taken for calculations $T_c$ | August 9-22 | October 1-14 | Nov 21-Dec 4 |
| $I_i$ | 7525 | 17605.4705407746 | 74507.6732607497 |
| $R_i$ | 73424 | 200051.672316368 | 548996.183882108 |
| $N_i$ | 336210.688000000 | 566602.33216 | 1454508.4416 |
| $\nu_i$ | 166678.304458998 | 271873.891638209 | 667978.033362204 |
| $\alpha_i$ | 6.47063569e-07 | 6.81143662034146e-07 | 2.13865037324172e-07 |
| $t_i^*$ | 201 | 255 | 306 |
| $\rho_i$ | 0.107851458558717 | 0.185185178161924 | 0.142857147036735 |
| $1/\rho_i$ | 9.27201183334553 | 5.40000020479830 | 6.99999979520000 |
| $r_i$ | 0.999591012582638 | 0.999637260478932 | 0.999333327224955 |
| $F_i$, eq. (23) | 14661.3785892930 | 16531.7953887678 | 8990.91853799112 |
| $F_i / F_C(1, n_i - 2)$ | 788.246160714677 | 888.806203697193 | 483.382717096297 |
| $S_{i\infty}$, eq. (11) | 91157 | 168 285 | 376 290 |
| $V_{i\infty}$, eq. (18) | 245054 | 398 317 | 1 078 218 |
| $t_{if}$, eq. (19) | 443.5 | 432 | 520 |
| Final day of the epidemic wave | April 8, 2021 | March 27, 2021 | June 23, 2021 |

Fig. 2 illustrates the COVID-19 pandemic waves in Ukraine calculated with the use of the optimal values of SIR parameters presented in Table 2 and formulas (10)-(17). Solid lines correspond to the numbers of victims *V=I+R*, dashed lines show the numbers of persons spreading the infection *I* multiplied by 10, dotted lines illustrate the theoretical estimations of the daily numbers of new cases *dV/dt* calculated with the use of eq. (14) and multiplied by 10. The red, black and blue colors correspond to sixth, seventh and eighth epidemic waves respectively. The accumulated number of cases *V*_*j*_ taken for calculations are shown by circles; “triangles” and “stars” correspond the cases taken only for comparisons and verifications of the predictions.

**Fig. 2.**
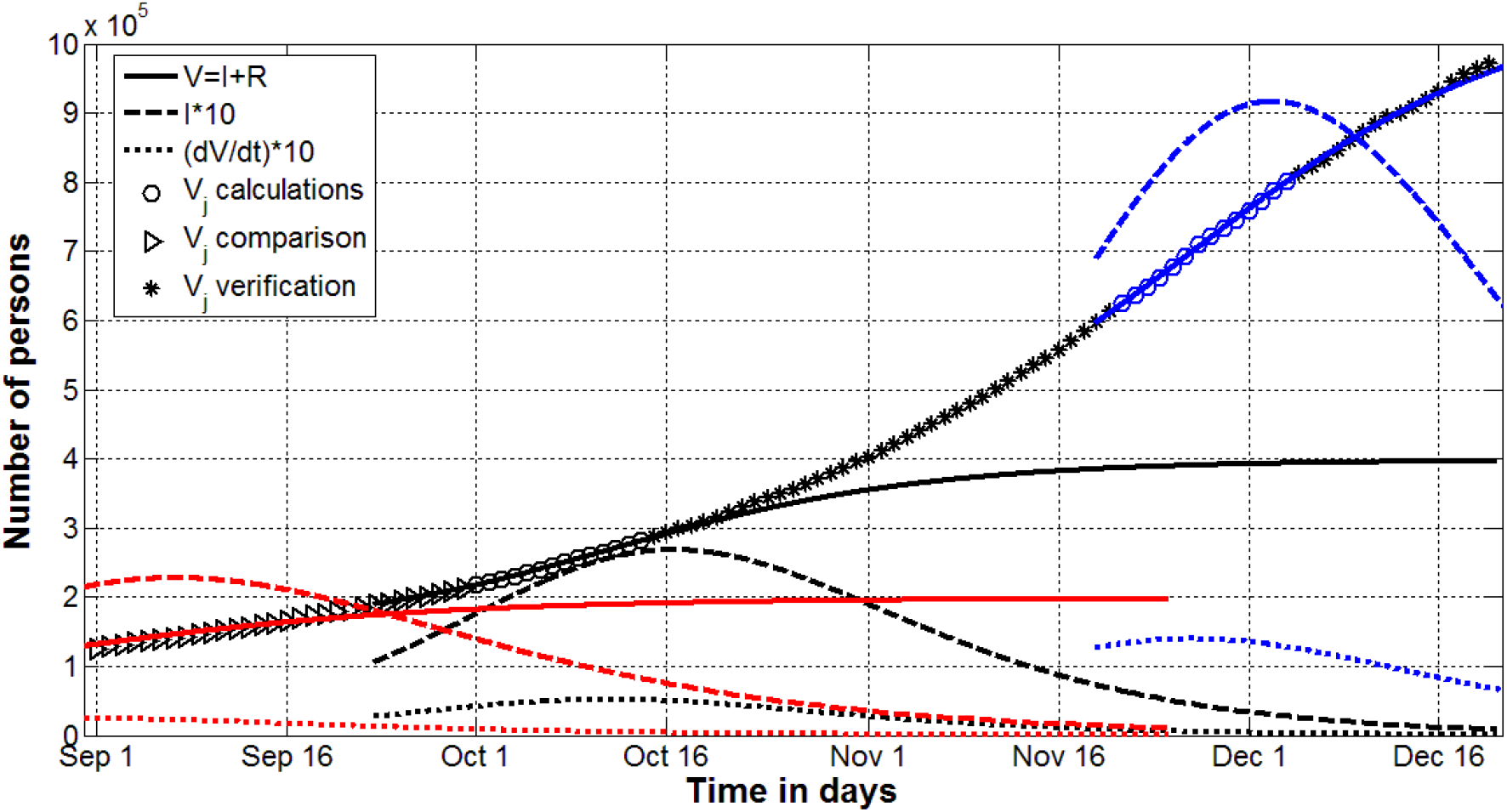
Sixth (red), seventh (black) and eighth (blue) COVID-19 pandemic waves in Ukraine in autumn 2020. Solid lines - the numbers of victims *V=I+R*, dashed lines - numbers of persons spreading the infection *I* multiplied by 10 infected, dotted lines - the theoretical estimations of the daily numbers of new cases *dV/dt* multiplied by 10. The accumulated number of cases *V*_*j*_ taken for calculations are shown by circles; “triangles” and “stars” correspond the cases taken only for comparisons and verifications of the predictions.

Fig. 2 illustrates rather good accuracy of SIR simulations. In particular, the *V*_*j*_ values (“triangles”) started to deviate from the solid red line only after September 16. This fact can be explained by the beginning classes in schools and universities (as mentioned above). This curve was calculated with the use of data set from the period August 9-22. The increased number of contacts in schools and universities caused rapid increase of the number of cases. This increase in the number of diseases intensified in October and November 2020 through elections and a presidential poll. The very irregular nature of the epidemic dynamics (particularly the large values of the second derivative (see “stars” in Fig. 1)) led to the fact that *V=I+R* curve for the seventh wave very quickly began to deviate from the recorded number of cases *V*_*j*_ (compare solid black line and “stars” in Fig.2).

The absence of sharp jumps of the second derivative after November 21, 2020 (see “stars” in Fig. 1), allowed us to predict quite accurately the further epidemic dynamics (compare solid blue line and “stars” in Fig.2). Blue dashed line in Fig.2 show that the number of persons spreading the infection diminished in December 2020. Let us hope that the Christmas and New Year celebrations will not significantly worsen this positive dynamics.

## Conclusions

Constant changes in the Covid-19 pandemic conditions (i.e., in the peculiarities of quarantine and its violation, in situations with testing and isolation of patients, in coronavirus activity due to its mutations etc.) lead to new pandemic waves and cause the changes in the values of parameters of the mathematical models. To identify these changes, a simple method was proposed based on the numerical differentiation of smoothed dependences of the accumulated number of cases. The results show that smoothing dependence of the accumulated number of cases and its differentiation can provide fairly accurate and useful information about the course of the epidemic, identify important changes in its dynamics and provide timely recommendations for quarantine measures or control of social distancing.

To simulate different pandemic waves (periods with more or less constant values of its dynamics parameters) a new generalized SIR model and its exact solution have been proposed. Procedure of its parameters identification were developed and successfully applied to calculate the characteristics of several pandemic waves in Ukraine.

The pandemic duration projections are unfortunately not optimistic. We hope that effective quarantine measures and successful vaccination will be able to reverse this trend. Very long duration of the pandemic requires correction of our behavior, we can not live as before it occurred. Decreased feelings of insecurity and non-compliance with social distancing may further increase the pandemic duration and the number of the coronavirus victims. Total closure of settlements or regions can be recommended only in the event of a sharp increase in the number of cases. There are many things that can be done without loss to the economy and our daily lives:

1. *Minimize the number of contacts and trips, not visit crowded places. Work and study remotely where possible*.
2. *Refrain from shaking hands and kisses during meetings. Use masks in transport and crowded areas*.
3. *If you (or others) have any suspicious symptoms, do your best to avoid the spread of the infection*.

## Data Availability

All data are in the text

## Acknowledgements

I would like to express my sincere thanks to Professors Dirk Langemann (Techniche Universitaet Braunschweig) and Juergen Prestin (Universitaet zu Luebeck) for their support in developing the used optimization approach. I would like to thank also Professors Alberto Redaelli, Giuseppe Passoni and Gianfranco Fiore (Politecnico di Milano), S. Pereverzyev (RICAM, Linz, Ausria) for involving me in very interesting biomedical investigations in frames of EU-financed Horizon-2020 projects EUMLS (Grant agreement PIRSES-GA-2011-295164-EUMLS) and AMMODIT (Grant Number MSCA-RISE 645672).

I would also like to thank Academician Viktor Grinchenko, Professors Volodymyr Tymofeyev, Alexander Galkin, Pavlo Maslianko; my friends Liudmyla Trotsenko, Maia Troian, Nina Basiuk, Anna Voronka, Daria Cusitcaia, Damien Berezenko, Anatolii Podkur, Volodymyr Borysenko and Oleksii Rodionov for their support and help in collecting and processing data.

